# Infective Endocarditis in Pediatric Patients with Cardiac Implantable Electronic Devices: A Survey of the Pediatric and Congenital Electrophysiology Society (PACES)

**DOI:** 10.64898/2026.02.05.26345706

**Authors:** Anjan S Batra, Morcel Hamidy, Anthony McCanta, Lauren Sell, Michael J Silka

## Abstract

**Background:** Guideline recommendations for infective endocarditis (IE) prophylaxis have narrowed significantly over the past decade. However, these recommendations are derived from adult data and may not adequately account for the unique risk factors for IE in pediatric and congenital heart disease (CHD) patients with cardiac implantable electronic devices (CIEDs).

**Objective:** To characterize contemporary IE cases and prophylaxis practices among members of the Pediatric and Congenital Electrophysiology Society (PACES) and assess how these practices align with or diverge from current international guidelines or practice recommendations.

**Methods:** A cross-sectional, web-based survey was distributed to PACES members worldwide. Questions addressed prophylaxis practices for CIED implantation, reinterventions, and bacteremia-inducing procedures, as well as clinician experience with IE in patients with CIED. Responses were analyzed descriptively.

**Results:** Substantial practice heterogeneity was identified across multiple clinical scenarios. Although most clinicians aligned with guideline recommendations for patients with structurally normal hearts, nearly all respondents (92.3%) reported recommending lifelong prophylaxis for patients with complex or repaired CHD. Among 35 reported IE cases, 97% occurred in transvenous systems, 77% occurred >6 months post-implantation, and 90% lacked a clear procedural or infectious trigger. Despite successful device extraction in 77% of cases, significant morbidity and mortality were observed.

**Conclusion:** Current practice patterns among pediatric and congenital electrophysiologists reflect uncertainty regarding the applicability of adult-derived IE prophylaxis guidelines to younger patients with CIEDs. High observed morbidity, long-term device exposure, and distinct anatomic considerations highlight the need for pediatric-specific risk stratification and updated guidance.

## Introduction

Infective endocarditis (IE) is a rare but potentially life-threatening complication in patients with cardiac implantable electronic devices (CIEDs), including transvenous pacemakers and implantable cardioverter-defibrillators (ICDs). Device-related infections can result in significant morbidity, including lead-associated endocarditis, sepsis, and the need for complete system extraction, which carries additional procedural risks (1–3). Although infection rates following CIED implantation in adults are generally low, pediatric and congenital heart disease (CHD) populations may have an elevated risk because of smaller patient size, repeated surgical and catheter-based interventions, indwelling prosthetic material, and unique anatomic substrates (4–7).

Over the past decade, recommendations for IE prophylaxis have narrowed considerably. The American Heart Association (AHA) and European Society of Cardiology (ESC) guidelines emphasize prophylaxis only for patients at the highest risk of adverse outcomes from IE—such as those with prosthetic cardiac valves, prior IE, or select uncorrected or partially repaired congenital heart lesions (8–10). For CIED procedures specifically, current guidelines recommend a single pre-procedural dose of antibiotics but do not support the use of routine postoperative antibiotics (8,9). Furthermore, the presence of a CIED alone is not considered an indication for IE prophylaxis during dental, gastrointestinal,genitourinary, or respiratory procedures (8,10).

However, these recommendations are derived almost exclusively from adult studies, with minimal pediatric-specific evidence regarding risk-benefit considerations. Children with CHD face a lifetime of increased IE susceptibility, more frequent exposure to bacteremia-provoking procedures, and higher prevalence of prosthetic material—factors that may place them at higher risk than the adult populations from which current guidelines were developed (4–7,11). CIED infections in pediatric patients can be especially consequential, given the challenges of vascular access preservation and the potential long-term implications of lead extraction or venous stenosis (12).

Despite these concerns, little is known about contemporary IE prophylaxis practices among pediatric and congenital electrophysiologists. Prior data suggest substantial heterogeneity in peri-procedural management, but practice patterns have not been comprehensively assessed since major updates to international guidelines. A 2018 PACES survey demonstrated wide variability in peri-operative antibiotic strategies and revealed that most pediatric electrophysiologists do **not** strictly follow adult-derived recommendations, often citing concerns about higher infection risk in children (13). Understanding current practice patterns is essential to identify gaps between guidelines and real-world care and to inform future pediatric-specific recommendations.

This study aimed to assess contemporary global practice patterns related to IE prophylaxis among members of the Pediatric and Congenital Electrophysiology Society (PACES). By evaluating prophylaxis strategies for CIED implantation, revision, and commonly encountered non-cardiac procedures, we sought to characterize current approaches, quantify areas of practice variation, and identify opportunities for standardization and further research.

## Methods

### Study Design

We performed a cross-sectional, descriptive study using an anonymous, web-based survey to evaluate contemporary infective endocarditis (IE) prophylaxis practices among clinicians caring for pediatric and congenital patients with cardiac implantable electronic devices (CIEDs). The study was designed to characterize real-world practice patterns and to capture clinician experience with device-related IE.

### Study Population

The survey was distributed electronically to the global membership of the Pediatric and Congenital Electrophysiology Society (PACES). Eligible respondents included physicians and advanced practice providers actively involved in the management of pediatric or adult congenital heart disease patients with CIEDs. Trainees were not specifically targeted.Participation was voluntary, and responses were collected anonymously. No patient-identifying information was obtained.

### Survey Development and Content

The survey instrument was developed by a multidisciplinary group of pediatric electrophysiologists with experience in device management and infectious complications. The questionnaire was pilot-tested by several PACES members for clarity and content validity prior to distribution. The final survey consisted of multiple-choice and categorical questions addressing the following domains:

1. **Respondent characteristics** – years in practice, professional role, and practice setting.
2. **Institutional CIED populations** – estimated numbers of patients followed with transvenous, epicardial, and leadless pacing systems.
3. **Antibiotic prophylaxis practices** – clinician-reported approaches to IE prophylaxis across a variety of clinical scenarios, including:
  - Routine dental procedures
  - Complex dental procedures
  - Minimally invasive gastrointestinal, genitourinary, or respiratory procedures
  - Body piercings
  - Peri-procedural management for new CIED implantation or reintervention
4. **Patient subgroups** – prophylaxis practices were queried separately for:
  - Patients with structurally normal hearts
  - Patients with repaired congenital heart disease at low risk of IE
  - Patients with repaired congenital heart disease considered at high risk of IE
  - Immunocompromised patients
5. **Device-specific considerations** – differences in practice for transvenous systems, epicardial systems, and leadless pacemakers.
6. **Clinical experience with infective endocarditis** – respondents were asked to report details of any cases of IE encountered in patients with CIEDs, including timing of infection relative to implantation, presence of preceding procedures, use of prophylactic antibiotics, management strategy, and outcomes.

### Definitions

For the purposes of the survey:

- **CIEDs** included transvenous pacemakers, implantable cardioverter-defibrillators (ICDs), and epicardial pacing systems. Insertable cardiac monitors were explicitly excluded.
- **High-risk congenital heart disease** was defined according to current AHA/ESC guideline categories, including patients with prior IE, unrepaired cyanotic heart disease, or repaired congenital heart disease with residual defects adjacent to prosthetic material.
- **Minimally invasive procedures** referred to gastrointestinal, genitourinary, and respiratory tract procedures with potential for transient bacteremia (e.g.,colonoscopy, bronchoscopy, cystoscopy).
- Reported IE cases were defined as clinician-diagnosed bacterial endocarditis involving a CIED system, managed at the respondent’s institution.

Respondents were asked to report cases and practices from the contemporary era, reflecting experiences and management patterns from 2010 through 2024.

### Survey Administration

The survey was hosted on a secure online platform and distributed via email to PACES members with an invitation letter describing the purpose of the study. Reminder emails were sent periodically over the study period to maximize participation. The survey remained open for responses for 10 weeks. Completion of the survey implied informed consent.

### Data Collection

All responses were automatically captured within the survey platform and exported for analysis. No personal identifiers or institutional identifiers were collected. Each respondent completed the survey once; duplicate entries were prevented by survey settings.

### Outcomes

The primary outcomes of interest were:

1. Reported patterns of IE prophylaxis across common clinical scenarios in pediatric and congenital patients with CIEDs.
2. Degree of concordance or discordance between clinician practice and current AHA/ESC guideline recommendations.
3. Descriptive characteristics of clinician-reported cases of CIED-associated IE, including timing, triggers, management, and outcomes.

### Statistical Analysis

Analyses were descriptive in nature. Categorical variables are presented as counts and percentages. No comparative hypothesis testing was planned a priori, as the primary objective of the study was to characterize practice variation rather than to evaluate specific predictors of behavior. Graphical summaries were generated to illustrate variation in prophylaxis strategies across clinical scenarios. All analyses were performed using standard statistical software (Microsoft Excel and SPSS version XX, IBM Corp., Armonk, NY).

### Ethical Considerations

Because the study involved anonymous survey responses from clinicians and did not include patient-level data, it was considered minimal risk. Institutional Review Board review was obtained at the coordinating institution, which determined that the study qualified for exemption.

## Results

### Respondent Characteristics

A total of 67 members of PACES completed the survey. The majority were physicians (63/67, 94.0%), with a smaller proportion of advanced practice providers (4/67, 6.0%). No respondents identified as trainees or selected “other.” The distribution of clinical experience was: 7 (10.4%) had <5 years in practice, 11 (16.4%) had 5–10 years, 24 (35.8%) had 10–20 years, and 25 (37.3%) had >20 years.

### Device Populations Managed

The number of CIED patients followed at respondents’ institutions varied widely. For transvenous devices, 40 of 66 respondents (60.6%) reported caring for >100 patients, while 14 (21%) reported 50–100 patients, 10 (15%) had 20–50 patients, and 2 (3.0%) had <20. For non-transvenous (epicardial) systems, centers reported a distribution: 22 of 66 (33.3%) followed 50–100 patients, 18 (27.3%) followed >100, 17 (25.8%) followed 20–50, and 9 (13.6%) followed <20. Leadless pacemaker experience was lower overall: among 65 respondents, most centers (51; 78.5%) followed <5 patients, while only 8 centers (12.3%) reported >10 leadless device patients.

### Prophylaxis Practices in Pediatric Patients with Structurally Normal Hearts (Figure 1)

For **routine dental procedures**, 38 of 65 respondents (58.5%) did not prescribe antibiotic prophylaxis, 23 (35.4%) recommended prophylaxis for the first 6 months post-implant, and 4 (6.2%) recommended prophylaxis indefinitely.

**Figure 1.**
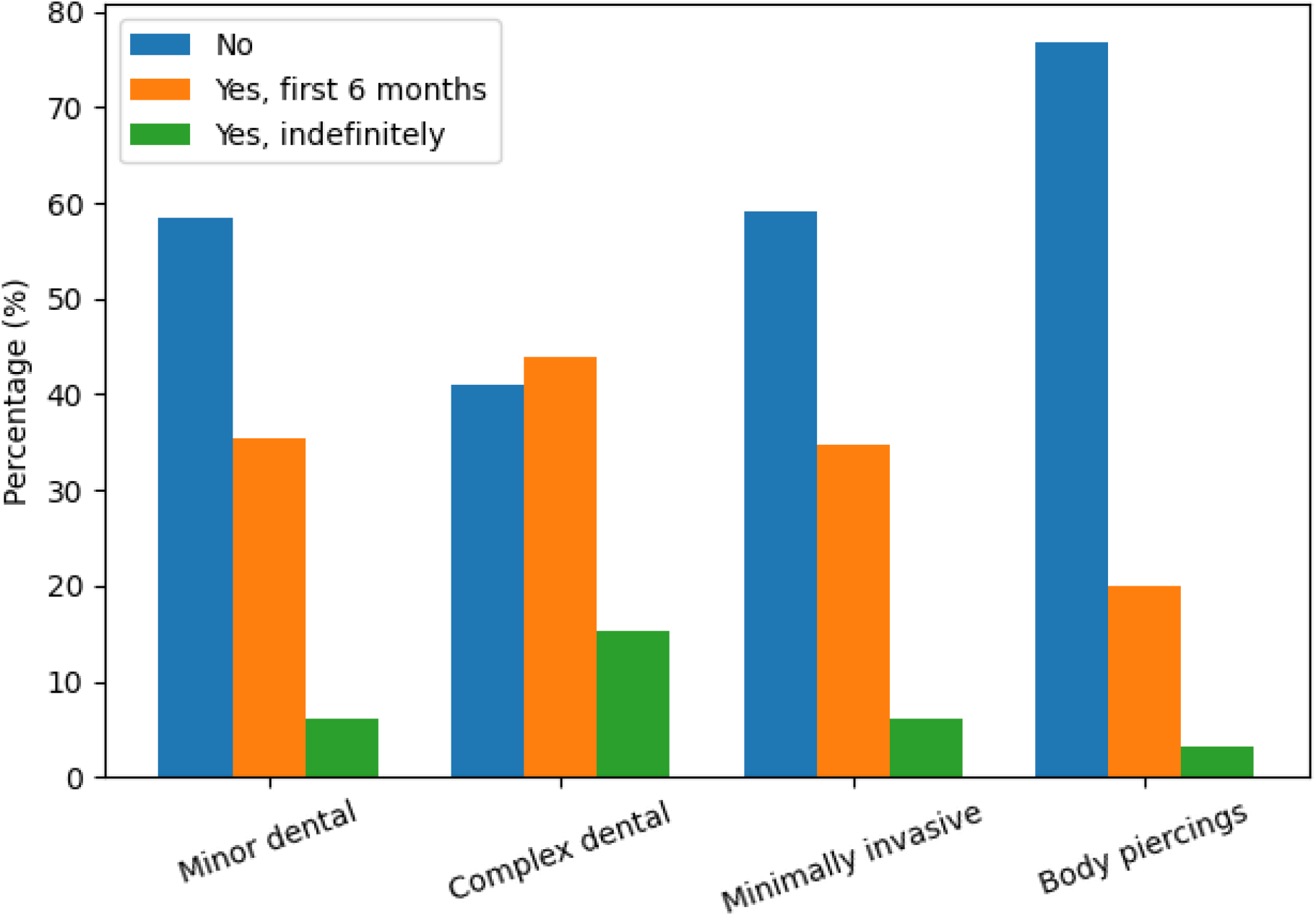
IE Prophylaxis Practices in CIED patients with Structurally Normal Heart. Each procedure category (minor dental, complex dental, minimally invasive procedures, and body piercings) is shown with three grouped bars representing: No prophylaxis, Yes, for the first 6 months post-implant and Yes, indefinitely.

For **complex dental procedures**, use of prophylaxis increased: 27 of 66 (40.9%) did not recommend prophylaxis, 29 (43.9%) endorsed prophylaxis for the first 6 months post-implant, and 10 (15.2%) endorsed indefinite prophylaxis.

Prophylaxis for **minimally invasive procedures** (colonoscopy, bronchoscopy, GU procedures) was similar to routine dental practices: 39 of 66 (59.1%) did not recommend prophylaxis, 23 (34.8%) recommended 6-month prophylaxis, and 4 (6.1%) recommended indefinite prophylaxis.

Most respondents (50 of 65, 76.9%) did not recommend prophylaxis for **body piercings**, though 13 (20.0%) endorsed 6-month prophylaxis and 2 (3.1%) endorsed indefinite prophylaxis.

### Prophylaxis in Pediatric Patients with Congenital Heart Disease (Figure 2)

For **routine or complex dental procedures** in patients with **repaired congenital heart defects at low risk of IE**, 22 of 66 (33.3%) recommended prophylaxis for the first 6 months post CIED implant, 5 (7.6%) recommended indefinite prophylaxis, and 39 (59.1%) did not prescribe routine prophylaxis.

**Figure 2.**
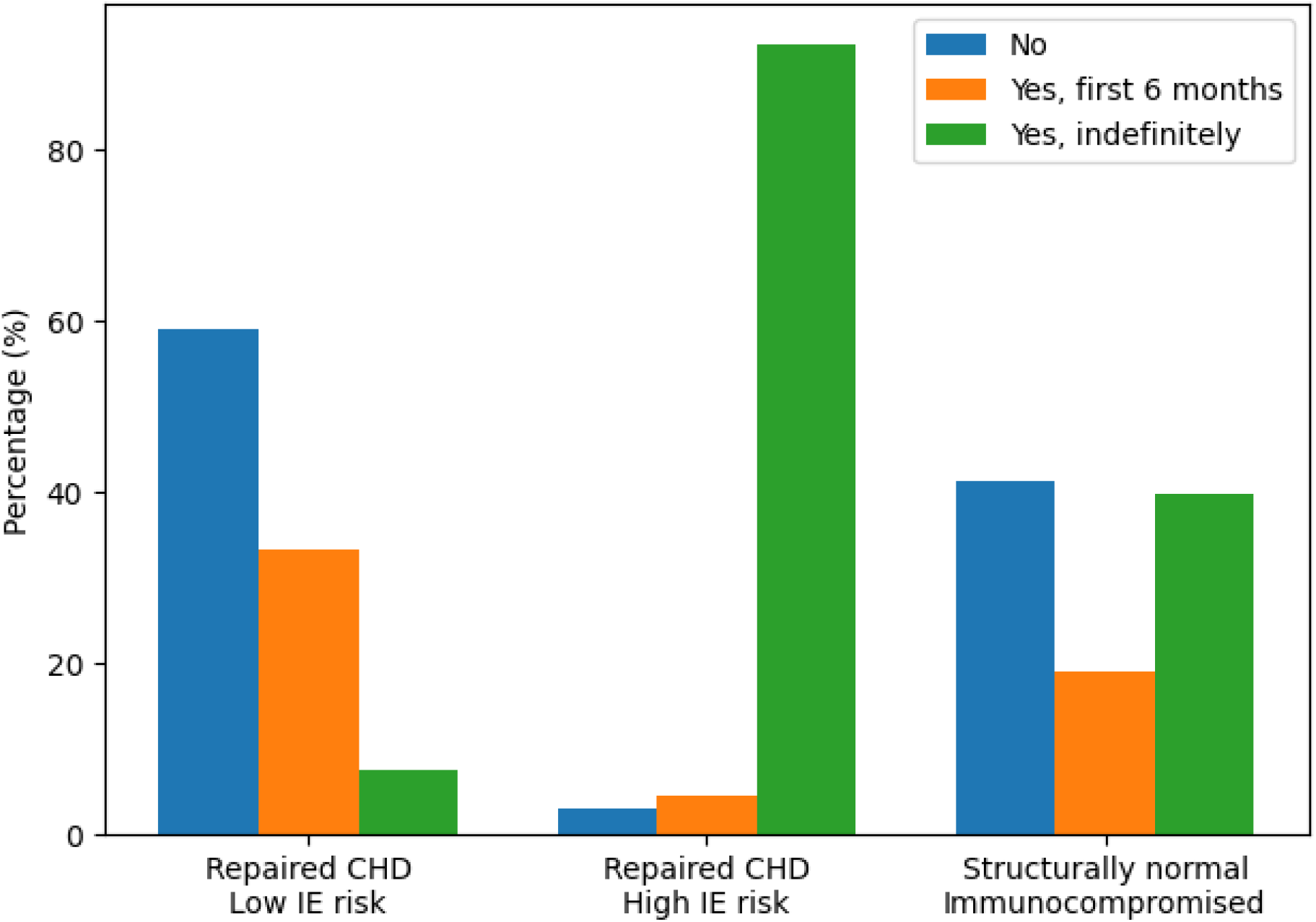
IE Prophylaxis Practices in CHD patients with CIED. Repaired CHD, low IE risk: Majority recommend no prophylaxis, with about one-third recommending limited (6-month) coverage. Repaired CHD, high IE risk (with prosthetic material): Overwhelming recommendation for indefinite prophylaxis. Structurally normal but immunocompromised: Recommendations are more evenly split, with substantial support for indefinite prophylaxis.

For **routine or complex dental procedures** in patients with **repaired congenital heart disease at high risk of endocarditis**, nearly all respondents (60 of 65; 92.3%) recommended **indefinite prophylaxis post CIED implant**, with only 3 (4.6%) endorsing 6-month prophylaxis and 2 (3.1%) not recommending prophylaxis.

For **immunocompromised patients with normal cardiac anatomy**, practice patterns were variable: 26 of 63 (41.3%) did not recommend prophylaxis, while 12 (19.0%) recommended 6-month and 25 (39.7%) recommended indefinite prophylaxis.

### Device-Specific Prophylaxis Practices

For **epicardial devices**, 47 of 67 respondents (70.1%) did not prescribe prophylaxis for dental or minimally invasive procedures. Eleven (16.4%) recommended prophylaxis for postoperative congenital heart disease with high IE risk, 6 (9.0%) recommended 6-month prophylaxis, and 3 (4.5%) recommended indefinite prophylaxis.

For **leadless pacemakers**, 30 of 63 (47.6%) did not recommend prophylaxis, 25 (39.7%) recommended 6-month prophylaxis, 7 (11.1%) recommended prophylaxis for high-risk congenital heart disease, and 1 (1.6%) endorsed indefinite prophylaxis.

### Antibiotic Selection

Among 64 respondents, nearly all (62; 96.9%) reported amoxicillin as their antibiotic of choice. Two respondents (3.1%) selected another agent.

### Incidence of Endocarditis

Respondents reported 35 cases of bacterial endocarditis, 34 of which occurred in patients with transvenous CIEDs. Most infections (27 cases; 77%) were diagnosed more than 6 months after device implantation (Figure 3). Postoperative congenital heart disease was present in 21 affected patients (60%). No preceding procedure was identified in 90% of cases, and prophylactic antibiotics had not been used prior to the onset of IE in 93%.Endocarditis resolved in 77% of patients following device lead extraction, although residual complications (n = 3) and deaths (n = 3) occurred.

**Figure 3.**
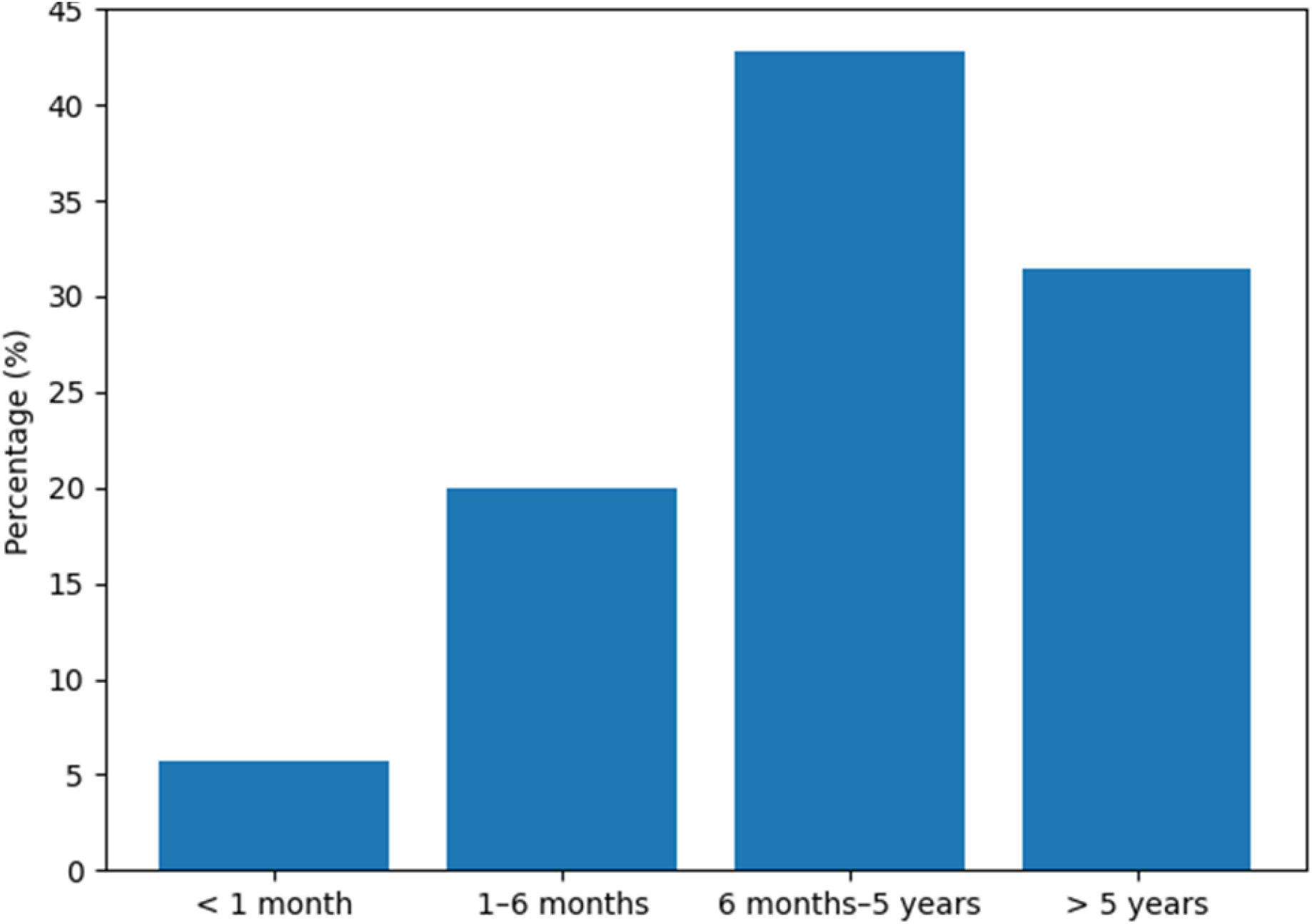
Interval Between CIED Implantation Diagnosis of IE. Most CIED-associated endocarditis events occurred beyond 6 months post-implant, with a clear peak in the 6 months–5 years interval, and nearly one-third occurring >5 years after implantation—highlighting the predominance of late infections.

## Discussion

In this international survey of members of the Pediatric and Congenital Electrophysiology Society (PACES), we identified substantial heterogeneity in clinical practice regarding infective endocarditis (IE) prophylaxis for pediatric and congenital patients with cardiac implantable electronic devices (CIEDs). Current guidelines from both the American Heart Association (AHA) and European Society of Cardiology (ESC) recommend IE prophylaxis only for a narrowly defined subset of cardiac conditions and do not include CIEDs as independent indications for prophylaxis.(8-10) However, our findings demonstrate that, in practice, clinicians caring for congenital and pediatric patients often adopt individualized, and sometimes divergent, approaches to antibiotic prophylaxis. This discrepancy highlights ongoing uncertainty regarding the applicability of existing guidelines to younger patients and those with congenital heart disease.

Guidelines restricting IE prophylaxis were originally shaped by adult data demonstrating low rates of procedure-related IE and concerns about antibiotic resistance and adverse drug reactions. (14,15) These guideline revisions significantly curtailed prophylaxis for routine dental, gastrointestinal, and genitourinary procedures. However, the adult populations in which these recommendations were validated differ substantially from pediatric and congenital patients who have complex anatomy, prior surgical repairs,indwelling prosthetic materials, and lifelong exposure to intravascular hardware. Pediatric and adult congenital heart disease (ACHD) patients with CIEDs may experience biomechanical and hemodynamic conditions that do not mirror those seen in adults receiving devices for age-related conduction disease. (12, 16, 17)

Importantly, transvenous leads in children traverse developing vasculature and may remain in place for decades rather than years. In children with CHD—particularly those with residual shunts, patches, conduits, or prosthetic material—the interaction between a chronically implanted lead and the vascular endothelium may create a unique substrate for infection. (17) These distinctions raise the possibility that adult-centric IE prophylaxis frameworks may not be applicable to pediatric and CHD populations.

Our survey captured 35 cases of bacterial endocarditis, nearly all (34 of 35) occurring in patients with transvenous devices. A striking observation was that 77% of infections occurred more than six months after implantation, and 90% occurred without a clear procedural trigger. These findings are consistent with broader data indicating that device-related IE in CIED recipients often occurs outside the peri-implantation period and tends to be driven by hematogenous seeding rather than procedural bacteremia. (1,18,19)

This challenges a central assumption underlying current prophylaxis recommendations— namely, that procedure-related bacteremia is a significant driver of IE in CIED recipients. If most infections arise spontaneously or from non-procedural bacteremia, then restricting prophylaxis solely to procedural events may fail to address the true mechanisms of infection in pediatric patients. Moreover, the high proportion of patients with postoperative CHD in our cohort further supports the notion that underlying anatomy and surgical history—rather than procedure type—drive risk in many pediatric cases.

Despite successful resolution in most affected patients following lead extraction (77%), morbidity and mortality remained significant, with three residual complications and three deaths reported. Prior studies in adults have documented that CIED-IE is associated with substantial morbidity and mortality: in one large prospective cohort, in-hospital mortality was 14.7% and 1-year mortality was 23.2%.(1) Another large series found that device removal improved survival compared with conservative treatment.(20) Moreover, pediatric and congenital patients face additional risks: venous access may be limited, vascular anatomy altered from prior surgeries, and lead extraction technically more challenging,increasing the risk of complications or long-term sequelae.(17,21)

These data emphasize that prevention — not merely treatment — should remain a central goal in this population.

Although most respondents did not routinely prescribe prophylaxis for patients with structurally normal hearts, nearly all (92.3%) recommended indefinite prophylaxis for patients with repaired CHD lesions considered high risk for IE. This suggests that clinicians are not disregarding guidelines but rather interpreting them in light of anatomic complexity, device type, and personal or institutional experience. In the absence of pediatric- or congenital-specific evidence, such variation is perhaps a rational response to uncertainty. Prior literature in congenital cardiology has documented similar inconsistencies in prophylaxis practices.(16,19)

Taken together, these findings highlight a critical gap in the evidence base guiding IE prophylaxis in pediatric and congenital patients with CIEDs. Adult-derived guidelines may underappreciate age-specific and anatomical risk factors, the long duration of device exposure in pediatric patients, and the cumulative lifetime risk of IE. As such, future studies are needed to define risk-stratification models that meaningfully incorporate:

- Congenital heart disease complexity
- Presence of residual or prosthetic cardiac material
- Lead type, age, and duration
- Vascular anatomy and prior interventions
- Frequency and type of bacteremia-inducing exposures

Multicenter, prospective registries and collaborative studies will be essential to determine true incidence, delineate procedural versus spontaneous infection triggers, and identify modifiable risk factors. Ultimately, such data may support tailored prophylaxis recommendations for pediatric and congenital patients with CIEDs, perhaps differing from adult guidelines.

### Limitations

This study has several limitations inherent to survey-based research. First, response bias may have influenced the results. The survey was completed by a subset of PACES members who may not be fully representative of all pediatric and congenital electrophysiology practitioners. Clinicians with stronger opinions or more frequent encounters with infective endocarditis (IE) may have been more likely to participate, potentially skewing reported practices and case experiences. Second, the survey relied on self-reported data, including recall of IE cases, clinical scenarios, and prophylaxis practices, all of which are subject to inaccuracies and incomplete reporting. Third, because the incidence and characteristics of IE were not obtained through a standardized registry, detailed information on patient anatomy, device type, comorbidities, microbiology, and management strategies could not be independently verified.

Additionally, the study did not capture the total number of CIED patients followed by respondents, precluding calculation of true IE incidence rates. The survey design also did not assess institutional protocols, which may differ from individual practice patterns and could influence clinical decision-making. Finally, the cross-sectional format captures practices at a single point in time and may not reflect evolving guidelines or recent changes in clinical practice. Despite these limitations, this study provides important initial insights into current practice variability and highlights the need for more robust, prospective data in pediatric and congenital populations with CIEDs.

## Conclusions

Survey results demonstrate limited adherence to current AHA and ESC guidelines for IE prophylaxis in pediatric and congenital patients with CIEDs. Most IE cases occurred late after implantation and without an identifiable preceding procedure, suggesting that existing guideline frameworks—largely derived from adult populations—may not be fully applicable to pediatric and adult congenital CIED recipients. These findings highlight the need for updated population-specific guidance regarding IE prophylaxis in this growing patient group.

## Data Availability

All data is available on RedCap through Children's Hospital of Orange County

## APPENDIX A. REDCap Survey Instrument: SBE (IE) Prophylaxis in Patients With Cardiac Implantable Electronic Devices (CIEDs)

1. What is your profession?
  ∘ Physician
  ∘ Advanced Practice Provider
  ∘ In Training
  ∘ Other (please specify)
2. How many years have you been in practice?
  ∘ < 5 years
  ∘ 5–10 years
  ∘ 10–20 years
  ∘ 20 years

### Section 2. Device Population at Respondent’s Institution

3. How many patients are currently followed in your institution with transvenous devices (pacemakers and ICDs)?
  ∘ < 20
  ∘ 20–50
  ∘ 50–100
  ∘ 100
4. How many patients are currently followed in your institution with non-transvenous (epicardial) devices (pacemakers and ICDs)?
  ∘ < 5
  ∘ 5–10
  ∘ 10
5. How many patients are currently followed in your institution with leadless pacemakers?
  ∘ < 5
  ∘ 5–10
  ∘ 10

### Section 3. IE Prophylaxis Practices by Clinical Scenario

For each of the following scenarios, respondents were asked: “Do you prescribe antibiotic prophylaxis?”

Response options:

- No
- Yes, for the first 6 months post-implant
- Yes, indefinitely

6. Structurally normal cardiac anatomy undergoing routine minor dental procedures (e.g., cleaning, adjustment of braces)
7. Structurally normal cardiac anatomy undergoing complex dental procedures (e.g., extractions, root canals, crowns)
8. Structurally normal cardiac anatomy undergoing minimally invasive procedures (e.g., colonoscopy, bronchoscopy, genitourinary procedures)
9. Structurally normal cardiac anatomy undergoing body piercings
10. “Repaired” congenital heart defects at low risk of endocarditis
11. “Repaired” congenital heart defects at high risk of endocarditis
12. Structurally normal cardiac anatomy in an immunocompromised patient

### Section 4. Device-Type–Specific Prophylaxis Practices

13. Do you routinely prescribe prophylactic antibiotics for patients undergoing dental or minimally invasive procedures post-implant for non-transvenous (epicardial) devices?
  - No
  - Yes, for the first 6 months post-implant
  - Yes, indefinitely
  - Yes, if they have postoperative congenital heart disease with high risk of endocarditis
14. Do you routinely prescribe prophylactic antibiotics for patients undergoing dental or minimally invasive procedures post-implant for leadless devices?
  - No
  - Yes, for the first 6 months post-implant
  - Yes, indefinitely
  - Yes, if they have postoperative congenital heart disease with high risk of endocarditis

### Section 5. Antibiotic Selection

15. What is your antibiotic of choice for SBE prophylaxis?
  - Amoxicillin
  - Other (please specify)

### Section 6. History of Endocarditis in Device Patients

16. Has endocarditis occurred in any of your patients with implantable devices?
  - Yes
  - No
17. If yes, how many cases?
  - 1
  - 2
  - 3
  - 4
  - 5 or more

### Section 7. Case-Specific Details for Each Reported Endocarditis Episode

For each episode (Case #1, Case #2, Case #3, etc.), respondents were asked:

a. Device type
  - Transvenous
  - Non-transvenous
  - Leadless
b. Time post-implant when endocarditis occurred
  - < 1 month
  - 1–6 months
  - 6 months–5 years
  - 5 years
c. Age of patient at time of endocarditis (Open-ended response)
d. Was there a procedure that may have led to endocarditis?
  - Dental
  - Other minimally invasive
  - No known procedure
e. If yes, how long before the onset of endocarditis did the procedure occur? (Open-ended response)
f. Were prophylactic antibiotics given prior to this procedure?
  - Yes
  - No
g. Outcome of endocarditis
  - Resolved with preservation of device
  - Resolved with removal of infected lead/device
  - Resolved but with residual complications
  - Fatal outcome
h. Did the patient have postoperative congenital heart disease?
  - Yes
  - No

